# A preliminary study to evaluate the exposure of human fetus to heavy metals in the umbilical cord blood in Syria

**DOI:** 10.1101/2024.03.17.24304428

**Authors:** Maryana Kabbaa, Ekbal Fadel, Flora Mayhoub

**Affiliations:** Department of Zoology, Faculty of Sciences, Tishreen University, Latakia, Syria; Department of Cytology, Embryology and Histology, Faculty of Medicine, Tishreen University, Latakia, Syria

**Keywords:** Human Fetus, Blood, Umbilical Cord, Heavy Metals, Lead, Mercury, Cadmium, Chromium, Nickel, Arsenic, Syria

## Abstract

The environmental situation in Syria needs a comprehensive assessment, especially in light of the conditions it has been experiencing for thirteen years, which have exacerbated pollution with heavy metals (HM) in various regions, including the coastal one.

This preliminary and first survey in Syria aims to evaluate the exposure of human fetus in the population to HM by measuring the toxic metals spread in the coastal environment in the umbilical cord blood (UCB), which are lead (Pb), mercury (Hg), cadmium (Cd), arsenic (As), chromium (Cr) and nickel (Ni).

The study was conducted between May 2022 and April 2023 among healthy newborns of the National Maternity Hospital in the Mediterranean coastal city of Tartous. This study adopted the official method of the American Association for Analytical Chemistry (AOAC, 2002) in collecting, preserving and processing UCB samples, and the heavy metals were measured using an Atomic Absorption Spectrometer. The statistical study was carried out using SPSS Statistics 23.0 (Statistical Package for Social Sciences).

The lower and upper limits for the concentrations of the studied elements in UCB range between: Pb (6.18-17.60µg/L), Hg (1.05-7.62µg/L), Cd (0.01-0.67µg/L), As (0.30-5.70µg/L), Cr (0.02-0.43µg/L), Ni (0.01-0.94µg/L).

The concentrations of all HM measured in UCB are below the recommended international reference limits. This paper represents the first step in studying the assessment of fetal exposure to HM in our region. The current and future studies aim to expand the study area to include all of Syria, in addition to linking laboratory levels of HM with various sources of exposure and pregnancy outcomes observed at birth.

## Introduction

Studying the exposure of the pregnant mother to heavy metals (HM) in particular is considered as very important due to the ability of these elements to cross the placental barrier and travel through the umbilical vein to spread throughout the fetus’s body and accumulate in its tissues and organs that are still forming and developing [1, 2].

There are many negative effects known so far on the health of the fetus, including immediate ones, such as prematurity, delayed growth, congenital malformations and intrauterine death [3], and long-term ones such as respiratory diseases, childhood cancers and others [4].

Evaluating the exposure of the pregnant mother and fetus to HM is of interest to many researchers in various countries around the world, as many studies have been conducted on the detection and calibration of HM in fetal biological matrices such as the placenta [5, 6, 7, 8], fetal membranes [8], umbilical cord tissues [5, 8, 9], umbilical cord blood (UCB) [6, 7, 9, 10, 11, 12, 13, 14, 15], amniotic fluid [16, 17, 18], neonatal hair [19, 20, 21], neonatal urine [10] and meconium [22].

In addition to the fact that these researches aim to study the negative effects of this exposure on fetal development in general [3, 7, 23, 24, 25], each of them seeks to establish an approved database in the country of study [4, 11, 15, 18, 26]. These studies pave also the way for subsequent researches in this field which means directing public and official opinions towards adopting the necessary means to reduce the levels of this exposure.

The environmental situation in Syria needs a comprehensive assessment, especially in light of the condition it has been going through for more than thirteen years, which affects various areas of life. The percentage of pollution with HM has increased because of the war in which various types of weapons were used, and the deterioration of the quality of petroleum derivatives spread in the markets. Weak control over industrial facilities, in addition to internal migration, led to an increase in residential clusters, industrial activities and traffics, especially coastal cities.

Many local studies conducted in the coastal region showed high levels of toxic HM such as lead (Pb), mercury (Hg), arsenic (As), chromium (Cr) and nickel (Ni) in the three components of the environment [27, 28, 29, 30, 31, 32, 33], and some elements of the food chain [34, 35, 36, 37, 38, 39].

As for epidemiological studies in humans in our country, they were limited to only two studies: it was observed in the first study [40] that higher concentrations of Pb and Ni in the hair of workers in cable, printing, and battery factories compared to workers in olive presses, textile and iron factories; the second study [41] showed a significant increase in the concentrations of Pb, Cd, and Ni in human hair for samples of workers in the field of mining, plastics and batteries compared to workers in administrative jobs.

Despite the large amount of local research concerned with environmental pollution with HM in our country, studies concerned with exposure of the general population, especially pregnant mothers and fetuses, are completely absent.

Based on the above and on our knowledge of the danger of fetal exposure to HM widespread in our country and our awareness of the necessity of evaluating this exposure, we decided to conduct this first survey of its kind in Syria to evaluate fetal exposure to toxic HM widespread in the local coastal environment and to create a database that will be a reference for future studies in the country.

After reviewing previous international studies aimed at evaluating fetal exposure to HM and their negative effects on the health of the fetus and child, and referring to local studies interested in evaluating environmental pollution with HM in the local environment, it was decided to detect the following elements in UCB: Pb, Hg, Cd, As, Cr and Ni.

## Materials and Methods

This study was extended over a full year, from May 2022 until April 2023. The study population consisted of newborns in the National Maternity Hospital in the city of Tartous (Figure 1). This hospital receives the highest number of annual births in the governorate (about 2,500 births/year) and from various areas of Tartous governorate, both rural and urban.

**Figure 1.**
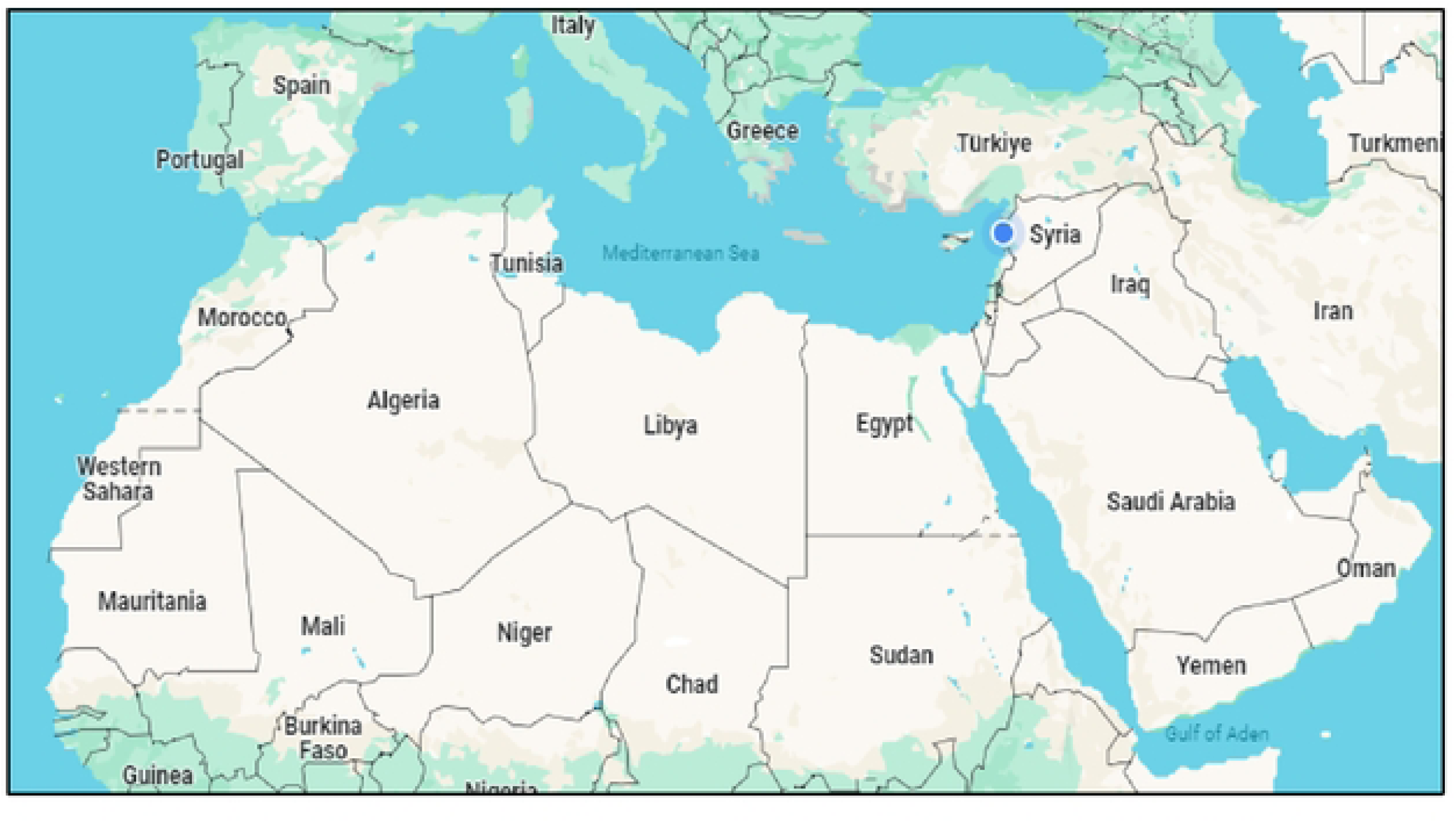
A map from Google Maps showing the location of the city and country of present study (Tartus•, Syria)

Before starting the study, approval was obtained from the Medical Research Ethics Committee at the Faculty of Human Medicine, certified by the Chairman of the Bioethics Committee at Tishreen University on March 31, 2022, in accordance with the ideal system of the Declaration of Helsinki regulating the ethics of medical research.

Before giving birth, the title of the study was presented verbally to the mother by the medical and nursing staff. In case of approval, the mother was given an informed consent document to sign. Upon signing, the newborn was given a special serial number for admission corresponding to the day of birth. The informed consent document initially included the title of the study, a simplified explanation of the subject of the study, then the nature of the required participation, emphasizing the absence of any interactions that could affect the health of the mother or the newborn, and the confidentiality of the data and results of the study participants, with the study team prepared to answer all questions and inquiries.

As a preliminary study aimed at investigating and revealing levels of exposure to HM in the general population, no exclusion criteria were set. Rather, all healthy neonates born in the National Maternity Hospital in the city of Tartous whose parents agreed to enter the study and signed informed consent were entered.

The general characteristics of the sample member’s studies in the current paper include maternal age and weight before pregnancy, maternal height, gestational age, and weight, length, and head circumference of the newborn at birth.

The official method of the American Association for Chemical Analysis (AOAC, 2002) was adopted in this study. Immediately after birth of the fetus and cutting of the umbilical cord and before the birth of the placenta, the midwife withdraws 5ml of umbilical vein blood from the piece connected to the placenta, using a syringe. The sample is stored in a 50ml conical plastic tube, 5ml of nitric acid is added to it, then the container is closed and numbered with the newborn’s serial number given upon entry into the study.

The tubes containing cord blood samples are collected and kept at room temperature in a designated place in the hospital and transported weekly to the scientific research laboratory at the Faculty of Sciences at Tishreen University to be processed. The control sample is prepared by gently heating 5ml of concentrated nitric acid and supplementing the volume of all samples to 25ml with distilled water. Samples preserved with nitric acid are heated in a water bath using an electric hotplate at a temperature of 100 degree Celsius for at least two hours. At the end of this stages, 2ml of hydrogen peroxide is added to each sample and heating is resumed for an additional hour until digestion is complete. The samples were then cooled and diluted with double-distilled water up to 50ml, where it becomes ready for calibration. It is then transferred to the Marine Chemistry Laboratory at the Higher Institute for Marine Research of Tishreen University to measure the HM using a Varian 220 Atomic Spectrometer Absorption device, according to the technologies specific to each element and according to the analytical conditions specific to each technology (Table 1).

The data obtained from this study were processed using the statistical program SPSS, 23.0 (Statistical Package for Social Sciences).

The statistical study was carried out using descriptive statistics measures (minimum and maximum, arithmetic mean (Standard Deviation (SD), in addition to the coefficient of variation (CV) and percentiles values) to study the distribution of levels of HM measured in UCB.

## Results

The total number of participants in this study was 210 couples (mother-newborn). For the mothers, the mean (SD) age was 28.2 years (5.8), the mean (SD) height was 1.63m (0.42), the mean (SD) weight before pregnancy was 62.4kg (5.6), and consequently the mean (SD) BMI before pregnancy was 23.4kg/m^2^. For the newborns, the percentage of males was 52.4%, the mean (SD) gestational age was 38.3 weeks (1.3weeks), the mean (SD) birth weight was 2974g (418.8), the mean (SD) birth newborn length was 51.3cm (51.3), the mean (SD) birth head circumference 33.8 cm (0.62).

Table 2 details the distribution of the HM concentrations in UCB studied in the present study. We note that it was possible to detect and quantify all the studied elements in the UCB samples. The values of UCB Pb concentrations range between (6.18-17.60µg/L). The median value of UCB Pb was 11.68µg/L. The mean (SD) value of UCB Pb was 11.67µg/L (2.76), and consequently the CV for UCB Pb concentrations is 24%. The values of UCB Hg concentrations range between (1.05-7.62µg/L). The median value of UCB Hg was 4.19µg/L. The mean (SD) value of UCB Hg was 4.06µg/L (1.50), and consequently the CV for UCB Hg concentrations is 37%. The values of UCB Cd concentrations range between (0.01-0.67µg/L). The median value of UCB Cd was 0.29µg/L. The mean (SD) value of UCB Cd was 0.31µg/L (0.15), and consequently the CV for UCB Cd concentrations is 48%. The values of UCB As concentrations range between (0.30-5.70 µg/L). The median value of UCB As was 2.33µg/L. The mean (SD) value of UCB As was 2.38µg/L (1.24), and consequently the CV for UCB As concentrations is 52%. The values of UCB Cr concentrations range between (0.02-0.43µg/L). The median value of UCB Cr was 0.19µg/L. The mean (SD) value of UCB Cr was 0.20 µg/L (0.10), and consequently the CV for UCB Cr concentrations is 50%. The values of UCB Ni concentrations range between (0.01-0.94µg/L). The median value of UCB Ni was 0.9µg/L. The mean (SD) value of UCB Ni was 0.09 µg/L (0.09), and consequently the CV for UCB Ni concentrations is 100%.

## Discussion

Available epidemiological studies interested in measuring HM concentrations in UCB adopt different descriptive statistical criteria, but most of these studies provide information about the lower and upper limits of their resulting concentration values. Therefore, we chose the upper limit criterion (Max) to compare with the results of international studies conducted in multiple countries spread over the five countries on one hand, and the range of values of the resulting concentrations (Min-Max) and the values of the Percentiles to compare with the range of the recommended international reference limits

Table 3 presents, in addition to our study, the similar international studies available to us. This table shows, for each study, the country, the year, the sample size (total and for each HM), the elements studied and their upper limits, and the range of international reference limits recommended for each of the elements studied. By regarding the data in tables 2 and 3, the following observations can be made:

1. **Lead (Pb):** In the current study, the descriptive statistical study of the values of UCB Pb concentrations shows clear closeness between these values for most of the samples. We note that the upper limit of the values of UCB Pb concentrations obtained in our study (17.60µg/L) is much lower than in other international studies with the exception of the study conducted in South Korea by Kim *et al.,* [42] in 2013 (15.8 µg/L). By comparing the range of the concentration values obtained in our study with the range of the international reference values (IRV) recommended not to be exceeded in the blood of children [43], which ranges between (35-50µg/L), we note that the values of UCB Pb concentrations in our study are less than the recommended limits in the blood of all samples.
2. **Mercury (Hg):** The descriptive statistical study of the values of UCB Hg concentrations resulting from our study indicates the distribution and divergence of the resulting values among them. The upper limit of the values of UCB Hg concentrations obtained in our study (7.62µg/L) is the lowest value among all the studies available to us. We note that the values of UCB Hg concentrations resulting in our study are less than the upper limit of the IRV recommended by WHO [44] not to be exceeded in the blood (3-10µg/L) for all individuals in the sample, it is even less than the lower limit for more than 25% of the sample (P25=2.95µg/L).
3. **Cadmium (Cd):** The value of the CV for UCB Cd concentrations in the current study (48%) indicates that the values of UCB Cd concentrations are far apart from each other, but by studying the rest of the statistical indicators, we notice that these values spread within a narrow range between (0.01-0.67µg/L). Although the upper limit values for UCB Cd concentrations resulting in our study (0.67µg/L) occupies the middle position among the available global studies; it remains the lowest at regional level, as a study conducted in Saudi Arabia by Al-Saleh *et al.* [45] (0.70µg/L), while a study conducted in Iran by Vigeh *et al.* [46] (6.3µg/L). On the other hand, we note that the values of UCB Cd concentrations resulting from our study are less than the IRV in the blood recommended by WHO [47] (<4 µg/L) for all individuals in the sample.
4. **Arsenic (As):** The descriptive statistical study of the values of UCB As concentrations resulting in our study indicates the large distribution of these values among them. When comparing the upper limit of the values of UCB As concentrations resulting in our study (5.70µg/L) with its equivalent in other international studies, we find it to be the lowest among these studies with the exception of the study conducted in South Africa by Röllin *et al.* [11] (2.84µg/L). We note that the values of UCB As concentrations obtained in our study are less than the IRV recommended in blood in the general population (<10µg/L) [48] in all participants in the study. To our knowledge, our study is the first of its kind in the Arab region and neighboring countries to evaluate As in UCB.
5. **Chromium (Cr):** The descriptive statistical study for the values of UCB Cr concentrations resulting from our study indicates a large distribution of these values among them, but they are all distributed within a narrow range ranging between (0.02-0.43µg/L). The upper limit of the values of UCB Cr concentration resulting in our study (0.43µg/L) is the lowest value among the available studies, including the study conducted in the neighboring country Turkey by Yüksel *et al.* [49] (1.20µg/L). We note that the values of UCB Cr concentrations are much lower than the lower limit of the range of IRV recommended not to be exceeded in the blood (20-30µg/L) [50] in all individuals participating in the study. To our knowledge, our study is the first of its kind in the Arab region to evaluate Cr in UCB.
6. **Nickel (Ni):** The values of CV (100%) and the Percentiles up to P95 for UCB Ni indicate that the values of UCB Ni concentrations obtained in our study are close to each other and distributed around the mean, with very few (<5%) outliers. Comparing the upper limit of the values of UCB Ni concentrations resulting in our study (0.94µg/L) with its counterpart in other available studies, we find that it is lower than in the Chinese studies conducted by Li *et al.* (21.2µg/L) [51] and the two Spanish studies conducted by Bocca *et al.* [19] and Dahiri *et al.* [52] (1.7 and 155.82 µg/L respectively), but higher than in the study conducted in the neighboring country Turkey by Yüksel *et al.* (0.59µg/L) [49]. We note that the values of UCB Ni concentrations obtained in our study are less than the lower limit of the range of IRV recommended not to be exceeded in the blood (1-4µg/L) [53] for all individuals in the studied sample. To our knowledge, our study is the first of its kind in the Arab region to evaluate Ni in UCB.

The values of Pb, Hg, Cd, As, Cr, Ni concentrations in UCB obtained by our study are among the lowest in the world, and they all located within the range of reference values recommended not to be exceeded in the blood by international organizations. The resulting values for Cr and Ni concentrations in UCB in our study clearly converge with their counterparts in the Turkish study, the only study completed on these two elements in UCB in the region.

It is difficult to find an explanation for these differences in the levels of heavy metals in UCB between our study and the others, but the spatial and temporal differences between these studies must have played a role in this. This is in addition to the diversity of sources and severity of pollution from one country to another. The difference in research methodologies and statistical study standards from one study to another makes it difficult to find uniform indicators between these studies so that researchers can collect the largest number of studies and determine the parameters of fetal exposure to heavy metals at local, regional, or international levels.

To our knowledge, our study is the first of its kind in the Arab region to evaluate As, Cr, and Ni, in human UCB.

## Conclusion

This paper represents the first step in studying the assessment of fetal exposure to heavy metals in Syria. Current and prospective studies conducted by our team to correlate laboratory levels of these elements with various sources of exposure and pregnancy outcomes observed at birth.

The results of this study reflect the state of fetal exposure to HM in the coastal Tartous Governorate, whose area and number of residents remain relatively small. Therefore, assessing this exposure requires a more comprehensive epidemiological survey that extends across multiple regions, especially with the geographical, climatic, and human diversity that characterizes our country.

## Data Availability

All relevant data are within the manuscript and its Supporting Information files.

## Supporting information

Table1 (DOCX)

Table2 (XLSX)

Table3 (DOCX)

## Acknowledgments

We thank the parents who participated in this study, the midwifery staff and administration at the National Maternity Hospital in Tartous, Higher Institute for Marine Research, Tishreen University for their contribution to completing this study.

We also thank Faculty of Sciences, Faculty of Medicine and the presidency of Tishreen University for their support to the success of this study.

Lastly, we thank Dr. Ahmed Kherbik for his help in data analysis and Pr. Amir Ibrahim for his help in editing this manuscript.

## Author Contributions

**- Conceptualization:** Ekbal Fadel, Flora Mayhoub

**- Methodology:** Flora Mayhoub

**- Validation:** Ekbal Fadel

**- Processing of UCB samples:** Maryana Kabbaa

**- Data curation:** Maryana Kabbaa, Flora Mayhoub.

**- Investigation:** Maryana Kabbaa, Flora Mayhoub

**- Supervision:** Ekbal Fadel, Flora Mayhoub

**- Writing - original draft:** Maryana Kabbaa

**- Writing - review & editing:** Ekbal Fadel, Flora Mayhoub

## References

1. Falcon M, Vinas P, Luna A. Placental lead and outcome of pregnancy. https://www.sciencedirect.com/journal/toxicology(2003), 10.1016/S0300-483X(02)00589-9.

2. Llanos MM, Ronco AM. Fetal growth restriction is related to placental levels of cadmium, lead and arsenic but not with antioxidant activities. Reprod Toxicol. 2009 Jan; 27(1):88±92. 10.1016/j.reprotox.2008.11.057 PMID: 19103280.

3. Caserta D, Graziano A, Lo Monte G, Bordi G, Moscarini M. Heavy metals and placental fetal-maternal barrier: a mini-review on the major concerns. European Review for Medical and Pharmacological Sciences, 2013; 17: 2198–2206.

4. Chen Z, Myers R, Wei T, Bind E, Kassim P, Wang G, et al. Placental transfer and concentrations of cadmium, mercury, lead, and selenium in mothers, newborns, and young children. J Expo Sci Environ Epidemiol. 2014 Sep-Oct; 24(5):537±44. 10.1038/jes.2014.26 PMID: 24756102.

5. Sakamoto M, Yasutake A, Domingo J, Chan H, Kubota, M, Murata K. Relationships between trace element concentrations in chorionic tissue of placenta and umbilical cord tissue: potential use as indicators for prenatal exposure. Environment International 60 (2013) 106–111, 10.1016/j.envint.2013.08.007

6. Al-Saleh I, Shinwari N, Mashhour A, Mohamed Gel D, Rabah A. Heavy metals (lead, cadmium and mercury) in maternal, cord blood and placenta of healthy women. Int J Hyg Environ Health. 2011 Mar; 214(2):79±101. 10.1016/j.ijheh.2010.10.001 PMID: 21093366

7. Iwai-Shimada M, Kameo S, Nakai K, Yaginuma-Sakurai K, Tatsuta N, Kurokawa N, et al. Exposure profile of mercury, lead, cadmium, arsenic, antimony, copper, selenium and zinc in maternal blood, cord blood and placenta: the Tohoku Study of Child Development in Japan. Environmental Health and Preventive Medicine (2019) 24:35 10.1186/s12199-019-0783-y

8. Kot K, Kosik-Bogacka D, Łanocha-Arendarczyk N, Malinowski W, Szymański S, Mularczyk M, et al. Interactions between 14 elements in the human placenta, fetal membrane and umbilical cord. Health, P. Int. J. Environ. Res. Public Health 2019, 16, 1615; doi:10.3390/ijerph16091615

9. Raghunath R, Tripathi R, Sastry V, Krishnamoorthy T. Heavy metals in maternal and cord blood. The Science of the Total Environment 250 2000 135–141. 10.1016/S0048-9697(00)00372-7

10. Odland JØ, Nieboer E, Romanova N, Thomassen, Y. Elements in placenta and pregnancy outcome in arctic and subarctic areas. International Journal of Circumpolar Health, 2004, 63:2, 169-187, DOI: 10.3402/ijch.v63i2.17703.

11. Röllin H, Rudge C, Thomassen Y, Mathee A, Odland JØ. Levels of toxic and essential metals in maternal and umbilical cord blood from selected areas of South Africa results of a pilot study. J. Environ. Monit., 2009, 11, 618–627, DOI: 10.1039/b816236k

12. Butler Walker J, Houseman J, Seddon L, McMullen E, Tofflemire K, Mills C, et al. Maternal and umbilical cord blood levels of mercury, lead, cadmium, and essential trace elements in Arctic Canada. Environ Res. 2006 Mar; 100(3): 295±318. 10.1016/j.envres.2005.05.006 PMID: 16081062

13. Grzesik-Gąsior J, Sawicki J, Pieczykolan A, Bień A. Content of selected heavy metals in the umbilical cord blood and anthropometric data of mothers and newborns in Poland: preliminary data. Scientific Reports (2023) 13:14077, 10.1038/s41598-023-41249-4

14. Gu T, Jia X, Shi H, Gong X, Ma J, Gan Z. An Evaluation of Exposure to 18 Toxic and/or Essential Trace Elements Exposure in Maternal and Cord Plasma during Pregnancy at Advanced Maternal Age. Int. J. Environ. Res. Public Health 2022, 19(21), 14485; 10.3390/ijerph192114485

15. Rahbar MH, Samms-Vaughan M, Dickerson AS, Hessabi M, Bressler J, Desai CC, et al. Concentration of lead, mercury, cadmium, aluminum, arsenic and manganese in umbilical cord blood of Jamaican newborns. Int J Environ Res Public Health. 2015 Apr 23; 12(5):4481±501. 10.3390/ijerph120504481 PMID: 25915835

16. Long M, Ghisari M, Kjeldsen L, Wielsøe M, Nørgaard-Pedersen B, Mortensen E, et al. autism spectrum disorders, endocrine disrupting compounds, and heavy metals in amniotic fluid: a case-control study. Molecular Autism (2019) 10:1 10.1186/s13229-018-0253-1

17. Maekawa R, Ito R, Iwasaki Y, Saito K, Akutsu K, Takatori S, et al. Evidence of exposure to chemicals and heavy metals during pregnancy in Japanese women. Reprod Med Biol. 2017; 16:337–348, DOI: 10.1002/rmb2.12049

18. Neamtu R, Craina M, Dahma G, Popescu A, Erimescu A, Citu I, et al. Heavy metal ion concentration in the amniotic fluid of preterm and term pregnancies from two cities with different industrial output. EXPERIMENTAL AND THERAPEUTIC MEDICINE 23: 111, 2022, DOI: 10.3892/etm.2021.11034

19. Bocca B, Ruggieri F, Pino A, Rovira J, Calamandrei G, Martínez M, et al. Human biomonitoring to evaluate exposure to toxic and essential trace elements during pregnancy. Part A. concentrations in maternal blood, urine and cord blood. Environ Res. July 2019; 177, 108599, 10.1016/j.envres.2019.108599

20. Baraquoni N, Qouta S, Vänskä M, Diab S, Punamäki R, Manduca, P. It takes time to unravel the ecology of war in Gaza, Palestine: Long-term changes in maternal, newborn and toddlers’ heavy metal loads, and infant and toddler developmental milestones in the aftermath of the 2014 military attacks. Int. J. Environ. Res. Public Health 2020, 17, 6698; doi:10.3390/ijerph17186698

21. Mohany K, El-Asheer O, Raheem, Y, Sayed A and El-Baz, M. Neonatal heavy metals levels are associated with the severity of neonatal respiratory distress syndrome: a case–control study. BMC Pediatr. 22, 635 (2022). 10.1186/s12887-022-03685-5.

22. Aziz S, Ahmed S, Karim S, Tayyab S and Shirazi A. Toxic metals in maternal blood, cord blood and meconium of newborn infants in Pakistan. EMHJ, Vol. 23 No. 10, 2017 23(10), 10.26719/2017.23.10.678

23. Garcia-Esquinas E, Perez-Gomez B, Fernandez-Navarro P. Lead, mercury and cadmium in umbilical cord blood and its association with parental epidemiological variables and birth factors. BMC Public Health 2013; 13:841–52. 10.1186/1471-2458-13-841 PMID: 24028648

24. Hu X, Zheng T, Cheng Y, Holford T, Lin S, Leaderer, B, et al. Distributions of heavy metals in maternal and cord blood and the association with infant birth weight in China. J Reprod Med. 2015; 60(1-2): 21–29, PMID: 25745747

25. Irwinda R, Wibowo N and Putri A. The concentration of micronutrients and heavy metals in maternal serum, placenta, and cord blood: a cross-sectional study in preterm birth. Hindawi Journal of Pregnancy Volume 2019, Article ID 5062365, 7 pages 10.1155/2019/5062365

26. Meyrueix L, Adair L, Norris S, Ideraabdullah F. Assessment of placental metal levels in a South African cohort. Environ Monit Assess. ; 2019, 191(8): 500. doi:10.1007/s10661-019-7638-2.

27. Kbibo I, Saker I, Ajeeb S. Study Of The Concentration Changes Of Some Heavy Metals In Chosen Sites From Elkabeer Northern River And Balloran Dam. Tishreen University Journal -Biological Sciences Series, 2001, 23(1). Retrieved from https://journal.tishreen.edu.sy/index.php/bioscnc/article/view/8215

28. Hammad Y, Mahmoud A. Study of some Bacteriological-Physico-Chemical Indicators and the Content of Some Heavy Metals in Alkabeer Alshamali River and two Neighboring Wells. Tishreen University Journal -Biological Sciences Series, 2010, 32(1). Retrieved from https://journal.tishreen.edu.sy/index.php/bioscnc/article/view/6062

29. Alia T, Nisafy I, Naseer R. A Case Study on the Impact of Human and Agriculture Activities on Some Drinking Water Quality Indicators in Qasmin Region. Tishreen University Journal -Biological Sciences Series,2012, 34(6). Retrieved from https://journal.tishreen.edu.sy/index.php/bioscnc/article/view/6311

30. Assad M, Abbas G, esmail M. Determination of some trace metals concentrations in storm water on Tartous previous coast. 2014 https://journal.tishreen.edu.sy/index.php/bassnc/article/view/788

31. Ghandour W, Laika H. Determination of some trace metals in the leaves and periderm of Avundodonax L. distribution in Lattakia city using Atomic Absorption Spectrophotmetry. Tishreen University Journal for Research and Scientific Studies-Biological Sciences Series Vol. (37) No (5) 2015.

32. Makhoul G, Ashee M, Alshouhna M, Khoury M. Studying the Effect of Tartous Cement Factory’s Dust on Content of Leaves for Khodairy Olive Var. from Some Heavy Metals. Tishreen University Journal -Biological Sciences Series, 2019, 41(1). Retrieved from https://journal.tishreen.edu.sy/index.php/bioscnc/article/view/7923

33. Shater Z, Ali W, Nesafi I, Saleh L. Biomonitoring Air Pollution Of Some Portable Heavy Metals in Dust Quarries By Using Needles Of Pinus brutia Ten, As Bio monitor In Site Of Kfardabeel Stand-Jableh. Tishreen University Journal -Biological Sciences Series, 2017, 39(1). Retrieved from https://journal.tishreen.edu.sy/index.php/bioscnc/article/view/3420

34. Kbebo E, Haifa S, Ziadeh M. Study Of Contents of the Soil, Surrounding Tartous Cement Plant of Some Heavy Metals (Pb, Cd, Fe, Cu, Zn, Ni). Tishreen University Journal -Biological Sciences Series, 2009, 31(5). Retrieved from https://journal.tishreen.edu.sy/index.php/bioscnc/article/view/6049

35. Mohamad I. A study of the Pollution of Some Syrian Coast Zones and Some Marine Organisms by Some Trace Heavy Metals. https://journal.tishreen.edu.sy/index.php/bassnc/article/view/5020

36. Mamish S. Study of jellyfish in Syrian Coastal water and their content of heavy and radioactive trace elements. Thesis Submitted in Partial of the Requirements for Master degree in Marine Biology, Department of Marine Biology, High institute of Marine research, Tishreen University, Lattakia, Syria P14, 2013.

37. Nouredden Se, Baker, M, Ammar I, Ali, A; Abbasse G, Abdo, O, et al. The Establishment of a regional network for tracing metals observation by biotical assembly in the eastern coast of the Mediterranean. Tishreen University Journal -Biological Sciences Series, 2012, 34(5). Retrieved from https://journal.tishreen.edu.sy/index.php/bioscnc/article/view/6288

38. Saker, Fayez; AL-Masri, Mohamad; Saleh, Mohamad. Trace Heavy Metals Accumulation in Some Zoo Benthic Species of Banias Littoral Thermal Station. Tishreen University Journal -Biological Sciences Series, 2008, 30(5). Retrieved from https://journal.tishreen.edu.sy/index.php/bioscnc/article/view/5964

39. Alnesser A, Abdullah, S. Study of accumulation of some heavy metals content in two species of algae (Jania rubens and Galaxura lapidescens) in the littoral zone of Lattakia. Tishreen University Journal -Biological Sciences Series, 2017, 39(4). Retrieved from https://journal.tishreen.edu.sy/index.php/bioscnc/article/view/4029.

40. Khuder A., Bakir M.A, Hasan R. et al. Determination of nickel, copper, zinc and lead in human scalp hair in Syrian occupationally exposed workers by total reflection X-ray fluorescence. Environ Monit Assess 143, 67–74 (2008). 10.1007/s10661-007-9958-x

41. Kherbik R. DETERMINATION OF SOME TRACE ELEMENTS IN HUMAN HAIR AS ENVIRONMENTAL POLLUTION INDICATOR. (2019). Iraqi Journal of Market Research and Consumer Protection, 11(1), 26–36. https://jmracpc.uobaghdad.edu.iq/index.php/IJMRCP/article/view/134

42. Kim YM, Chung JY, An HS, Park SY, Kim BG, Bae JW, et al. Biomonitoring of Lead, Cadmium, Total Mercury, and Methylmercury Levels in Maternal Blood and in Umbilical Cord Blood at Birth in South Korea. Int J Environ Res Public Health. 2015 Oct 26; 12(10):13482±93. 10.3390/ijerph121013482 PMID: 26516876

43. CDC. 2021. Preventing lead poisoning in young children. Atlanta, GA: US Department of Health and Human Services, Public Health Service, Centers for Disease Control.

44. Mercury, W. I. (1991). Environmental Health Criteria 118. Geneva: World Health Organization, 107.

45. Al-Saleh I, Shinwari N, Mashhour A, Rabah A. Birth outcome measures and maternal exposure to heavy metals (lead, cadmium and mercury) in Saudi Arabian population. Int J Hyg Environ Health. 2014 Mar; 217(2±3):205±18. 10.1016/j.ijheh.2013.04.009 PMID: 23735463

46. Vigeh M, Yokoyama K, Ramezanzadeh F, Dahaghin M, Sakai T, Morita, Y, et al. Lead and other trace metals in preeclampsia: a case– control study in Tehran, Iran. Environmental Research 100 (2006) 268– 275, doi: 10.1016/j.envres.2005.05.005

47. Fielberg L, Elinder C. in Cadmium, World health Organization, Geneva, 1992, Environmental Health Criteria, 134, page 17ff.

48. Agency for Toxic Substances and Disease Registry. 2007

49. Yüksel B, Arıca E, Söylemezoğlu T. Assessing reference levels of nickel and chromium in cord blood, maternal blood and placenta specimens from Ankara, Turkey. J Turk Ger Gynecol Assoc 2021; 22: 187–95, DOI: 10.4274/jtgga.galenos.2021.2020.0202.

50. Agency for Toxic Substances and Disease Registry (2000). “Toxicological Profile for Chromium.” http://www.atsdr.cdc.gov/toprofiles/tp7.html.

51. Li A, Zhuang T, Shi J, Liang Y, Song M. Heavy metals in maternal and cord blood in Beijing and their efficiency of placental transfer. Journal of Environmental Sciences, 10.1016/j.jes.2018.11.004

52. Dahiri B, Hinojosa M, Carbonero-Aguilar P, Cerrillos L, Ostos R, Bautista J, et al. Assessment of the oxidative status in mother-child couples from Seville (Spain): A prospective cohort study. 207, 308–319. https://www.sciencedirect.com/journal/free-radical-biology-and-medicine, 10.1016/j.freeradbiomed.2023.08.017

53. Base de données Biotox, sur le site web de I’INRS : www.inrs.fr/biotox-05/2023.

54. Al-Jawadi A, Al-Mola Z,Al-Jomard R.Determinants of maternal and umbilical blood lead levels: a cross-sectional study, Mosul, Iraq. licensee BioMed Central Ltd, doi:10.1186/1756-0500-2-47

55. El Khaleegy H, El-moghazy M, Abo baraka W. Study of Umbilical Blood Lead Level and Its Relation with Pregnancy Outcome. Journal of Medical and Pharmaceutical Sciences – AJSRP – Issue (2), Vol. (3) – June 2019, DOI : 10.26389/AJSRP.W040319

56. Alemam H, Enattah N, Fellah A, Elftisi E, Akarem A, Bashein A. Correlation between maternal and fetal umbilical cord blood lead concentrations in Libya. East Mediterr Health J. 2022;28(5):345–351. 10.26719/emhj.22.020

57. Kim J, Kim S, Woo S, Chung J, Hong Y, Oh S, et al. Prenatal exposure to lead and chromium is associated with IL-13 levels in umbilical cord blood and severity of atopic dermatitis: COCOA study. Immune Netw. 2019 Dec;19(6): e42 10.4110/in.2019.19.e42 pISSN 1598-2629·eISSN 2092-6685

58. Koppen G, Den Hond E, Nelen V, Mieroop E, Bruckers L, Bilau M, et al. Organochlorine and heavy metals in newborns: results from the Flemish Environment and Health Survey (FLEHS 2002–2006). Environment International 35 (2009) 1015–1022, doi: 10.1016/j.envint.2009.05.002

59. Saoudi A, Dereumeaux C, Goria S, Berat B, Brunel S, Pecheux M, et al. Prenatal exposure to lead in France: Cord-blood levels and associated factors: Results from the perinatal component of the French Longitudinal Study since Childhood (Elfe). International Journal of Hygiene and Environmental Health (2018), 10.1016/j.ijheh.2018.01.007.

60. Abdelouahab N, Huel G, Suvorov, A, Foliguet B, Goua V, Debotte, G, et al. Monoamine oxidase activity in placenta in relation to manganese, cadmium, lead, and mercury at delivery. teratology. Neurotoxicol. Teratol., 2009, 32 (2), pp.256–61. ff10.1016/j.ntt.2009.08.010ff.ffinserm-00422145f.

61. Yazbeck C, Cheymol J, Dandres A, Barbéry-Courcoux A. Lead exposure in pregnant women and newborns: a screening update. Elsevier Masson SAS. Tous droits réservés. doi:10.1016/j.arcped.2006.09.016

62. Guy M, Accrombessi M, Fievet N, Yovo E, Massougbodji A, Le Bot B, et al. Toxics (Pb, Cd) and trace elements (Zn, Cu, Mn) in women during pregnancy and at delivery, South Benin, 2014–2015. Environ Res. June 2018; 167, 198–206, 10.1016/j.envres.2018.06.054.

63. Barbieri F, Gardon J, Ruiz-Castell M, Paco V P, Muckelbauer R, Casiot C, et al. Toxic trace elements in maternal and cord blood and social determinants in a Bolivian mining city. International Journal of Environmental Health Research, 2016 Vol. 26, No. 2, 158–174, 10.1080/09603123.2015.1061114.

64. Téllez-Rojo M, Bautista-Arredondo L, Rosa-Parra A and Silva G. Prenatal exposure to metals and concentration thereof in umbilical cord blood in a Mexico City cohort. Gaceta Médica de México. 2023; 159 159(2), 132-137, DOI:10.24875/GMM.M23000759

65. Zheng G, Zhong H, Guo Z, et al. Levels of Heavy Metals and Trace Elements in Umbilical Cord Blood and the Risk of Adverse Pregnancy Outcomes: A Population-Based Study. Biol Trace Elem Res 160, 437– 444 (2014). 10.1007/s12011-014-0057-x

